# An exploration of the unmet needs of patients diagnosed with Idiopathic Pulmonary Fibrosis (IPF): a scoping review protocol

**DOI:** 10.1101/2022.11.25.22282749

**Authors:** Carita Bramhill, Donna Langan, Helen Mulryan, Jessica Eustace-Cook, Anne-Marie Russell, Anne-Marie Brady

## Abstract

**Introduction:** Interstitial lung diseases (ILDs) consist of a range of lung disorders with the most prevalent being Idiopathic pulmonary fibrosis (IPF)^1 2^. Idiopathic pulmonary fibrosis (IPF) is a chronic, progressive disease, resulting in loss of lung function and potentially significant Impacts on quality of life^1^. There is an increasing need to address unmet needs in this population as there is evidence that unmet needs may impact quality of life and health outcomes. The key objective of this scoping is to define the unmet needs of patients living with a diagnosis of IPF and to identify gaps in the literature relating to unmet needs. Findings will inform the development of services and the introduction of patient-centred clinical care guidelines for Idiopathic Pulmonary Fibrosis (IPF).

**Methods and Analysis:** This scoping review is guided by the methodological framework for conducting scoping reviews developed by the Joanna Briggs Institute^3^. The Preferred Reporting Items for Systematic Reviews and Meta-Analyses extension for Scoping Reviews checklist is used for guidance. The following databases will be searched CINAHL, MEDLINE, PsyhcoInfo (EBSCO platform), Web of Science (Core Collection), Embase (Elsevier), ASSIA Proquest). A comprehensive review of grey literature will be completed. Two independent reviewers will screen articles in consecutive stages from title/abstract screening to full-text screening for relevance against the inclusion and exclusion criteria. Data will be extracted using a predefined data extraction form. Data will be analysed using descriptive and thematic analysis. Findings will be presented in a general descriptive overview and tabular summaries coupled with a narrative summary of findings.

**Ethics and dissemination:** Ethics approval is not required for this scoping review protocol. This protocol was peer reviewed by Academics as part of the doctoral programme of study. We will disseminate our findings using traditional approaches that include open access peer-reviewed publication and scientific presentations.

**Strengths and Limitations of the study:**

➢ A comprehensive peer reviewed search strategy will be used to guide this review, incorporating expertise from multiple disciplines to maximize the effectiveness of the search strategy.
➢ This review will provide a comprehensive mapping of the literature related to Idiopathic Pulmonary Fibrosis (IPF) and unmet needs including a comprehensive grey literature search.
➢ This scoping review will apply the Joanna Briggs Institute (JBI) scoping review methodological framework guided by the Preferred Reporting Items for Systematic Reviews and Meta-Analyses extension for Scoping Reviews checklist.
➢ A team-based analysis will enhance analytical rigour and integrity of the findings.
➢ The full text review will limit to articles from 2011 to present, as anti-fibrotic medication for IPF was not widely available prior to 2011.
➢ Studies in languages other than English are excluded.

## INTRODUCTION

### Background

Interstitial lung disease (ILD) describes a group of heterogeneous diseases that are identified by inflammation and fibrosis of the lung interstitium^4 5^. A large subset of patients who are diagnosed with ILD have pulmonary fibrosis (PF). Most types of pulmonary fibrosis are identified by a progressive phenotype characterised by breathlessness, cough and fatigue^1 6^. The most common PF is IPF, and accounts for around 17-37% of all ILDs^7^. Idiopathic pulmonary fibrosis (IPF) is a chronic progressive disease with potentially enormous impacts on both physical and emotional well-being^8-10^

IPF is identified by irreversible loss of lung function with limited proven treatment options resulting in a life limiting condition coupled with impaired quality of life^11 12^. IPF typically occurs in older adults and is characterised by several disabling symptoms, including progressive worsening dyspnoea, impaired lung function, and for some patient’s anxiety and depression, carrying with it a poor prognosis^13 14 15^. The disease course can be varied from one patient to another. Some patients may experience slow disease progression and others may encounter periods of stability and finally there are some patients who experience a rapid decline or a disease trajectory with acute exacerbations^16^. An IPF diagnosis can have functional, psychological, and social impacts on individuals who benefit from an integrated care approach across the continuum from diagnosis and along the disease course^17^. Most patients with IPF die from respiratory failure within three to five years of diagnosis, making it a devastating disease^18-20^.

Patients diagnosed with IPF, have an uncertain prognosis, high symptom burden and may suffer from complications related to multiple potential comorbidities including other pulmonary conditions resulting in impaired quality of life^21 22^. In many cases, patients also require complex respiratory care particularly at the end of life^23^.

The mean age of IPF patients ranges from 65–70 years, with incidence increasing with age and with higher rates seen in males than females^4 24 25 26^. The numbers of patients with IPF are rising globally, possibly due to several factors including an aging population, more awareness of the disease and more advanced diagnostic tools^19^. Maher and colleagues estimate the adjusted incidence and prevalence of IPF to be in the range of 0.09-1.30 and 0.33-4.51 per 10,000 persons, respectively^27^.

Clinical management of IPF patients includes referral of patients for lung transplantation assessment, ^28 29^ pulmonary rehabilitation^1 30^ and supplemental oxygen^31^ and palliative care input^20 32^. A cure for IPF does not currently exist, although there are two approved drugs, pirfenidone and ninetedanib that may slow disease progression^1 33-35^. In recognition of the availability of pharmacological treatments, international IPF guidelines including the European IPF patient charter further emphasise the importance of early diagnosis and access to essential care and services^36^. The arrival of anti-fibrotic drugs signified a major improvement in patient care, with several studies qualifying the impacts of both. However, despite the advances in current treatments there is minimal evidence, regarding impacts on quality of life^12 37 38^.

In line with emerging evidence on the pathogenesis of the disease, knowledge is evolving regarding the impact of IPF on patients’ lives including not only the physical needs of patients but also the psychosocial needs of patients and carers^39^. IPF patient experiences and consequent needs are broad and can vary along the clinical course of the disease. It is understood that many of the anticipated needs of patients diagnosed with IPF remain challenging, with many unmet needs persistently not being met^8 40 41^. A diagnosis of IPF can carry with it a marked socioeconomic burden and financial strain on patients and their families reflected by the negative effects on quality of life, healthcare utilization and the ability to work^10^.

An initiative by 11 European patient advocacy groups for pulmonary fibrosis identified five key unmet needs which formed the basis for the European IPF Patient Charter and further qualified in numerous other studies: 1) Early and accurate diagnosis, 2) Early access to care, including medication and transplantation irrespective of age 3) A holistic approach to standardise IPF management 4) Comprehensive and high quality information about IPF and 5) Better access to palliative and end-of-life care^1 36^ (see figure 1 in supplementary material). It is understood that many of these needs remain unmet for this patient group. There is an urgent necessity to quantify patients’ unmet needs to benchmark standards against International best practice clinical guidelines and consolidate the network of specialist centres delivering patient centred care. Better understanding of IPF patients lived experience and comprehensively recording their needs will enhance the patient care pathway^30 42^.

The literature widely reports continuing delays in’ early diagnosis and referrals to specialist centres^6 10^. According to Van der Sar^6^ some of the delays relating to diagnosis relate to the heterogeneity of the disease, rarity of pulmonary fibrosis as well as the requirements for multiple investigations resulting in a prolonged delay in diagnosis^43^.

Non-Pharmacological treatment options, such as pulmonary rehabilitation, oxygen therapy, psychological support, lung transplantation and access to a specialist ILD nurse are a vital part of holistic care for patients with IPF^36 44 45^. Previous studies have demonstrated that non-pharmacological treatment options are not equally available for patients in different European countries^1 36^. There exists a need for relevant up to date information about IPF, more education and continuous counselling, to adequately support patients along the disease course^43 44^

Palliative care referral and a symptom-based approach which compliment disease-focused care is one of the key recommendations in the European IPF patient charter, yet most patients with IPF receive limited palliative care throughout the course of their disease or at the end of life^46 36^.

### Rationale

A scoping review is considered the most appropriate method in this case to map the available evidence, identify gaps in knowledge and guide potential research in a comprehensive and systematic way^3^. This evidence synthesis will outline the characteristics of the unmet needs of patients diagnosed with IPF. It will aim to outline some of the barriers and facilitators outlined in the literature in meeting patient’s needs.

This review is guided by a central question, which is to map the available evidence related to the unmet needs of patients living with a diagnosis of IPF. For the purpose of this review, patient needs will be mapped to the care needs identified by the European IPF patient charter in addition to emerging needs that may be identified in the literature, and to date have not been included in the European Patient Charter^36^.

A preliminary review of the literature (Medline and CINAHL conducted in May 2022) did not reveal any existing scoping reviews of patient’s unmet needs using a mixed methods approach. Much of the available research took place pre-covid-19 and did not consider a hybrid care model, for example a mix of telephone clinics coupled with face-to-face appointments. There also seems to be a sparsity of research reflecting IPF patients’ needs in the community. This is particularly significant as we move toward a model of community-based care across several international healthcare systems.

The mapping of the evidence will benefit planning for future care resources including an IPF clinical care pathway. This proposed scoping review will utilise JBI methodology and will highlight the available evidence and identify any deficiencies in the sources^3 47^. A preliminary search of JBI evidence synthesis, the Cochrane database of systematic reviews and Prospero was conducted prior to this protocol development, and no current or in-progress systematic or scoping reviews on the topic were identified.

A scoping review approach can offer several important features including providing a broad overview of the landscape in relation to available literature and is therefore the most appropriate design for this evidence synthesis ^48^.

### Review Question

The aim of this scoping review is to identify what evidence exists about the unmet needs of patients living with a diagnosis of IPF.

## Objectives

The objectives of this scoping review are:

1. To synthesise the unmet needs of patients living with a diagnosis of IPF.
2. Define barriers and facilitators to meeting patient’s needs.
3. Provide an overview of relevant concepts and terminology.

## Methods and Analysis

This scoping review will be conducted in accordance with the Joanna Briggs Institute framework for scoping reviews^3^ and includes the following steps: (i) identifying the research question, (ii) developing a search strategy, (iii) study selection and (iv) data analysis and presentation. The Preferred Reporting Items for Systematic Reviews and Meta-Analyses (PRISMA-ScR) checklist will also be used to guide the reporting quality of this study. This review will utilise the PCC framework (population, concepts and contexts) for this scoping review (Appendix four) ^3^.

### Inclusion Criteria

#### Types of Participants

This scoping review will focus primarily on literature reporting on adult patients >18 with a diagnosis of IPF. Studies that refer to patients with a diagnosis of pulmonary fibrosis will be included, owing to the similarities in the disease course between IPF and some forms of Pulmonary Fibrosis.

### Concept

The main concept in this review are the unmet needs of patients living with a diagnosis of IPF throughout the disease course. Selected studies will be required to include information on unmet needs such as information needs, digital literacy needs related to home monitoring for example, diagnostic needs, psychological needs, access to healthcare services and personnel, or needs related to access to pharmacological treatments and/or oxygen, or physical health needs.

### Context

This review will consider studies on unmet needs in all settings, such as clinics, hospitals, long term care centres, community centres, respiratory community hubs, ambulatory care, home and remote access services such as telemedicine, face -to -face outpatient appointments and virtual clinics either phone consultations or using technology such as Zoom video calls or a mix of both. There will be no geographic limits in this review. The review will be limited to publications in or after 2011 when anti-fibrotic treatment for IPF notably became available in Europe in 2011.

### Types of Information Sources

This review will consider studies that describe the unmet needs of patients living with a diagnosis of IPF and will also include studies that reference pulmonary fibrosis. We will include all review types including systematic, scoping and literature reviews which describe unmet needs of our patient group. This review will not include case reports, protocols, letters or posters, commentaries, and opinion pieces. This review will include sources that relate to patients with a diagnosis of pulmonary fibrosis, but these sources must also refer to patients with Idiopathic Pulmonary Fibrosis

### Search Strategy

The search strategy for this review was developed with the assistance of a JBI-trained medical research librarian and peer reviewed by experts in the field of chronic disease, specifically IPF. The search strategy was externally peer-reviewed by a second librarian using the Peer Review of Electronic Search Strategies (PRESS) guidelines^49^. First a limited search of MEDLINE (EBSCO) and CINAHL (EBSCO) was undertaken to identify information resources in this review. The text words contained in the titles and abstracts of relevant articles, and the index terms used to describe the articles, will be used to develop a full search strategy for Medline (EBSCO) and CINAHL (EBSCO), and will not apply limits. In the second stage, the Medline (EBSCO) search strategy, including all identified keywords and index terms, will be adapted, and tailored for each subsequent information source (see appendix one). Through this iterative process, new search terms may be identified and utilized. There will be no date or language restrictions applied at this initial stage.

The search will be extended beyond Medline (EBSCO) to the other databases identified for inclusion in this review APA PsycINFO, CINAHL, (EBSCO platform), Embase (Elsevier), and Web of Science (Core Collection) and ASSIA (Applied Social Science Index) (Proquest), to search for studies that meet the review’s inclusion criteria. These databases have been selected for their combined ability to generate a wide range of evidence, specific to the research topic, across a range of interdisciplinary fields. There will be no date limits used in the initial screening criteria, instead databases will be searched from inception to present. Date limit criteria will be applied at full text review (2011-present). The search will be limited to human participants over 18 years of age. A copy of the search strategy is available in the supplemental material (see appendix one).

Grey literature and unpublished studies will be included, sources include ProQuest Dissertations and Thesis Global, Google Scholar and ClinicalTrials.gov, WHO International, Clinical Trials Registry Platform and OpenGrey. There will also be a manual search of the reference lists of included studies, to identify studies that may have been missed within the initial search. Annual conference abstracts from relevant international conferences will also be reviewed.

### Source of Evidence Selection

All identified records will be collated and uploaded onto EndNote X9.3.3 (Clarivate Analytics, PA, USA) and duplicates removed. All identified citations will be transferred into Covidence software (Veritas Health Innovation, Melbourne, Australia) where any remaining duplicates will be removed.

Once the records have been transferred to Covidence software, the titles, and abstracts will be screened by two independent reviewers for assessment against the inclusion criteria. The criteria will be pilot tested on a sample of search results to ensure ‘substantial’ inter-rater reliability (ie. Kappa statistic > 0.61) among screeners^50^. Potentially relevant studies which meet the inclusion criteria will be retrieved in full text and uploaded to Covidence. Additional exclusion criteria may be applied to the full text review, due to the iterative nature of scoping reviews ^3^.

The full text of these selected citations will be assessed in detail against the inclusion criteria by two independent reviewers (PCC inclusion criteria). Resources that do not meet the inclusion criteria will be recorded and reported in the scoping review and the reason for their exclusion provided. If disagreements arise about the inclusion or exclusion of a paper, then the reviewers will discuss in detail so that the issue can be resolved. A third reviewer will arbitrate in such instances.

### Data Extraction

Data will be extracted from articles and other evidence sources included in the scoping review by two independent reviewers, using a data extraction tool developed by the study’s research team in adherence with the review objectives (see appendix two). The design of the instrument for data extraction follows the guidelines from the JBI. A number of specific domains will be included in the data capture tool including but not limited to, study characteristics such as author, year of publication, geographic location, context (where the study was conducted), status of the publication (eg, published or grey literature), journal, aims/purpose, sample characteristics (study population, sample size (if applicable) and key findings that relate to the scoping review question (specific needs categories) study aims, methodology/methods (study design and measures and analysis approach) and the study findings (eg, quotes, themes, concepts and study results) and limitations.

The draft data extraction tool will be modified and revised as necessary during a pilot testing phase prior to reviewing all articles independently. Data extraction will be done electronically. Modifications will be detailed in the full scoping review report. Any disagreements that arise between the reviewers will be resolved through discussion, or with a third reviewer. Authors of papers will be contacted to request missing or additional data, where necessary.

### Analysis of the evidence

#### Data presentation

Data analysis will involve descriptive quantitative (i.e., frequency analysis) and qualitative (ie, thematic) analysis. An overview of the literature will be undertaken through a descriptive numerical summary (eg, characteristics of included studies, types of study design, characteristics of the study population and geographical location, study aims, methodology and limitations) and through emerging themes (deductive thematic analysis). The extracted data will be presented in tabular and graphic form in a manner that aligns with the study’s objectives. A narrative summary will accompany the tabulated results, describing how the study’s findings relate to the review’s objectives and research question (Appendix three).

### Patient and Public Involvement

Patient and public representatives (n= 5) from the Irish Lung Fibrosis Association PPI group were involved in reviewing the research protocol. PPI involvement enables translation of knowledge into practice by disseminating results among potential knowledge users. The stakeholders comprise of patient representatives, family members of patients diagnosed with IPF and experts in the field of IPF. This iterative process will continue to seek stakeholders’ perspective on data screening, data extraction and analysis phases. The PPI meetings will continue to take place through videoconference to allow wider participation. All members of the PPI group have read and acknowledged the groups terms of reference. The authors anticipate continued work with the stakeholder group to provide ongoing consultation and feedback relating to dissemination of the evidence synthesis.

## Ethics and Dissemination

Ethical Approval for the conduct of this scoping review, essentially a secondary analysis of existing literature is not required. We aim to engage with our PPI partners to rigorously thread peer review throughout the development of the protocol and brought through various aspects of the scoping review. We will disseminate the findings to a wide range of stakeholders including researchers, clinicians, policy makers and health service administrators, patient groups and lay and public audiences, to widen participation. We will disseminate the findings in academic peer reviewed journal publications and local, national, and international conference presentations and social media platforms.

## Discussion

The primary purpose of this evidence synthesis is to identify the breath and scope of the literature relating to unmet needs for patients diagnosed with IPF. This research will aim to identify gaps in the literature and to provide a backdrop for future research focus regarding the unmet needs of patients diagnosed with IPF, as no scoping review of evidence relating to IPF patients, in a covid-19 era currently exists. The findings from this review could enhance service provision for patients living with IPF, based on the unmet needs identified in this review. The findings may also help guide policy makers when developing clinical care programmes in respiratory disease to consider the specific needs of patients diagnosed with Idiopathic Pulmonary Fibrosis.

In relation to the search strategy although it is comprehensive there is a risk that we may still miss potentially relevant literature. To overcome this limitation, we have recruited an experienced health science librarian to help us develop the search strategy and to engage in discussions to refine and amend, the search strategy to maximise its reach. In addition, as scoping review studies include a diverse range of literature including grey literature there are potential practical concerns regarding time, funding, staffing and access to resources which should be considered when planning a scoping review.

## Supporting information

Search Strategy

## Data Availability

Date relates to review of existing literature.

## Acknowledgement Section

With special thanks to the participants of the Irish lung Fibrosis Association PPI group.

