## Supplementary material for "An exploration of the unmet needs of patients diagnosed with Idiopathic Pulmonary Fibrosis (IPF): a scoping review protocol": Search Strategy

**Appendix 1 Search Strategy**

**Medline EBSCO**

Search conducted on 14^th^ May 2022

| **Search** | **Query** | **Records Retrieved** |
| --- | --- | --- |
| S1 | TI ( “Idiopathic pulmonary fibros*” OR “Idiopathic interstitial pneumonia*” OR “Familial Idiopathic Pulmonary Fibrosis*” OR “Usual Interstitial Pneumon*” OR “fibrosing interstitial lung disease” OR “progressive fibrosis” OR “nonspecific interstitial pneumonia” OR "pulmonary fibros*" ) OR AB ( “Idiopathic pulmonary fibros*” OR “Idiopathic interstitial pneumonia*” OR “Familial Idiopathic Pulmonary Fibrosis*” OR “Usual Interstitial Pneumon*” OR “fibrosing interstitial lung disease” OR “progressive fibrosis” OR “nonspecific interstitial pneumonia” OR "pulmonary fibros*") | 23,354 |
| S2 | (MH "Idiopathic Pulmonary Fibrosis") OR (MH "Idiopathic Interstitial Pneumonias+") OR (MH "Pulmonary Fibrosis+") | 26,014 |
| S3 | S1 OR S2 | 35,660 |
| S4 | TI ( (servic* OR need* OR support* OR care* OR caring OR nurs* OR pathway*) N4 (access* OR barrier* OR disparit* OR demand* OR gap* OR unmet* OR lack* OR challenge* OR obstacle* OR limit* OR difficult* OR capacit* OR burden* OR benefit*) ) OR AB ( (servic* OR need* OR support* OR care* OR caring OR nurs* OR pathway*) N4 (access* OR barrier* OR disparit* OR demand* OR gap* OR unmet* OR lack* OR challenge* OR obstacle* OR limit* OR difficult* OR capacit* OR burden* OR benefit*)) ) | 411,640 |
| S5 | (MH "Health Services Needs and Demand+") OR (MH "Health Services Accessibility") OR (MH "Healthcare Disparities") | 150,631 |
| S6 | S4 OR S5 | 516,375 |
| S7 | S3 AND S6 | 356 |

**Appendix 2: Data Extraction Instrument**

| **Evidence source details and characteristics** |
| --- |
| Author |
| Year |
| Title |
| Country |
| Aims/Purpose |
| Study Population and sample size |
| Concept |
| Context or setting |
| Study Design |
| Methodology |
| Patient needs identified (physical, psychological, other) |
| PROMS Utilised |
| Summary of key findings |
| Limitations |

**Appendix 3: Data Presentation example**

| **Paper Characteristic** | **Results** |
| --- | --- |
| Country | By country of origin |
| Year Published | By year |
| Journal | By journal |
| Study Design | 1.Randomised controlled trials.  2. Non-Randomised controlled trials.  3. Quasi-experimental studies.  4. Before and after studies  5. Prospective cohort studies.  6. Retrospective cohort studies  7. Case-control studies.  8. Cross sectional studies.  9. Other Quantitative studies.  10. Qualitative studies. |
| Patients’ Needs | 1. Early Diagnosis 2. Information needs 3. Access to medications & Oxygen 4. Access to pulmonary Rehabilitation. 5. Access to lung transplant. 6. Access to psychological support and palliative care. 7. Other |
| Population | 1. IPF 2. Pulmonary Fibrosis 3. ILD |

| **Framework** | **Element** | **Key Terms** |
| --- | --- | --- |
| **PCC** | **Population** | Adult patients over the age of 18 years who have a diagnosis of idiopathic Pulmonary Fibrosis. It will also include articles that refer to patients with a diagnosis of Pulmonary Fibrosis. |
|  | **Concept** | Healthcare needs. |
|  | **Context** | All care settings. |

**Appendix 4:**

**Table 1: PCC Framework for search strategy development**
